# Development of Explainable Machine Learning Framework for Early Detection and Risk Stratification of Diabetes in Age Specific Variations

**DOI:** 10.64898/2026.04.25.26351733

**Authors:** Nosizo Lukhele, Fahad Mostafa

## Abstract

**Objective:** To develop and evaluate a novel machine learning (ML) framework tailored to a clinical diabetes dataset and to assess whether demographic stratification enhances model performance and interpretability for multiclass diabetes classification.

**Methods:** A clinical dataset of 264 patients’ records was used to classify individuals into non-diabetic, prediabetic and diabetic categories. Several supervised learning models were trained using 80:20 train-test split and optimized using RandomizedSearchCV Model and 10-fold cross validation. Model performance was evaluated using the metrics accuracy, precision, recall and the F1-score. Area under the receiver operating characteristic curve (AUC) was calculated for the best generalizing model. A structured ML framework was developed for this dataset, incorporating preprocessing, model optimization, age stratification analysis age (<35 vs ≥35 years) and gender. SHAP was developed for model interpretability.

**Results:** Ensemble methods demonstrated superior performance in comparison to linear or single-tree approaches, with Gradient Boosting showing the most stable generalization with a test accuracy of 0.981 and stable cross validation accuracy of 0.972. AUC-ROC analysis using Gradient Boosting yielded good discriminative ability across the three diabetes classes: 0.991 (non-diabetic), 0.986 (prediabetic) and 0.972 (diabetic). Stratified analysis showed improved reliability in individuals aged ≥35 years (accuracy = 0.94, F1-score = 0.92), while performance in younger individuals was unstable due to small sample size. SHAP analysis identified HbA1c, BMI, and age as dominant predictors.

**Conclusion:** This study presents a ML framework integrating age stratified modelling with explainable ML frameworks to improve interpretability. The findings offer clinically relevant results that can support clinical decision-making systems, individualized risk assessment, and potential applications for targeted intervention in diabetes progression.

## Introduction

Diabetes Mellitus is a chronic metabolic disorder characterized by high blood glucose levels (hyperglycemia) due the underutilization and overproduction of glucose [1]. According to the International Diabetes Federation Diabetes Atlas, In 2019 it was reported that about 50% of people with diabetes were unaware that they have the disease, and by 2030, the prevalence is projected to increase to approximately 10.2% (578 million) [2]. Similarly; the World Health Organization predicted that diabetes would affect around 425 million people between the ages 20-79 years in 2023 and this figure is projected to increase to 629 million by 2045 [3]. The reason people go unaware that they have the disease or are at risk is because diabetes is largely asymptomatic and people do not preventatively screen for it until it has advanced. Delayed diagnosis of diabetes can lead to many preventable complications such as retinopathy, neuropathy, heart attacks and even strokes [4]. This makes diabetes a global concern, warranting the need for early detection and accurate diagnosis stage classification.

Diabetic risk is influenced by a combination of factors including age, BMI and biochemical factors. However, these clinical and sociodemographic factors are also shaped by genetics, lifestyle habits and one’s environmental exposures [5]. As a result, sole reliance on clinical thresholds is limiting and might not fully represent the dynamic as well as progressive nature of metabolic dysregulation [6]. This dependence on conventional clinical thresholds may result in underdiagnosis and delayed detection of individuals in intermediate stages, particularly those with prediabetes, reducing opportunity for early intervention [7]. Talaschian et al. (2026) showed that HbA1c screening varies by age and sex, with younger adults especially women requiring lower diagnostic thresholds for accurate detection [8], highlighting the limitations of universal diagnostic thresholds. As a result, clinicians need advanced data driven approaches and new technologies in order to deliver personalized diagnostic assessment and provide targeted intervention Machine learning tools offer a promising solution by integrating multidimensional clinical data to uncover complex patterns that traditional methods (like standardized questionnaires) cannot capture, thereby enhancing the accuracy of non-diabetic, pre-diabetic and diabetes prediction [9]. Research that supports the reliability of machine learning models for diabetes risk classification [10-11]. For example, Zhao et al. (2025) showed that ML models can achieve strong predictive performance for the progression of gestational diabetes to type 2 diabetes [12], demonstrating real-world applicability across diverse cohorts. Aragão et al (2025) evaluated different binary, multiclass and multilabel classification tools and found that they delivered consistent strong performance when applied in real world problems [13]. These works highlight how much ML yields improved predictive accuracy due to advancements in their computational power and algorithm complexities.

Despite the strong demonstrated performance, several research gaps remain. First, many of the existing ML studies frame diabetes prediction as a binary classification task (diabetic vs non-diabetic), which oversimplifies the disease since it progresses along a continuum. Prediabetes in particular is a very clinically meaningful stage where early intervention is most effective, hence it is important to classify this stage in modelling. Second, most ML frameworks apply a single universal model across heterogeneous populations, with limited investigation into whether model performance varies across demographic subgroups such as age and gender, which may limit model generalizability across different subgroups. To address these limitations, this study contributes to literature by integrating three complementary components within a unified analytical framework to show that machine learning can support clinicians with timely insights that enable clinicians to tailor treatments strategies.

In this paper, we have added the following contributions.

- Demographic-stratified modeling framework: This study contributes to literature by proposing a novel machine learning pipeline that evaluates age-based strata (<35 vs ≥35 years). This approach is grounded in updated screening recommendations from the US Preventive Service Task Force (USPSTF) and the American Diabetes Association (ADA), which lowered the diabetes screening age to 35 years to promote earlier detection and treatment [14]. This moves beyond applying universal models across heterogeneous populations and enables the uncovering of disease patterns distinct to specific subgroups or people.
- Development of Multiclass diabetes stage prediction: Unlike most ML approaches that simplify diabetes prediction into binary classification, this work models diabetes into three stages (non-diabetic, prediabetic, diabetic), providing a more clinically relevant representation of the disease.
- Development of explainable artificial intelligence Integration: By including SHAP within the framework, will enhance the interpretability and applicability of the ML models towards disease risk minimization [15-16].
- Identification of stage and subgroup-specific risk patterns: The integration of stratification and explainability will reveal how the importance of key predictors varies across age groups. This enables a more precise characterization of age-dependent risk profiles and supports more personalized risk assessment strategies.

These contributions greatly advance the modelling methodology of diabetes prediction and most importantly supports clinical decision making by demonstrating that interpretable, data-driven ML models can reliably enhance disease diagnosis and prognosis [17].

The article is organized as follows. The first section is the Introduction section: It introduces the clinical context, motivation and key research objectives. The second section: Literature Review examines existing works on machine learning approaches for diabetes prediction, highlighting current limitations. Third section: Methods describe the dataset, preprocessing steps, model development including the demographic stratification, evaluation frameworks and explainable AI technique used. The Fourth section: the Results present the performance of the ML models, subgroup analyses and SHAP- interpretations. Finally, the fifth section: Discussion & Conclusions summarizes the major findings and outlines what the implications are for public health and proposes what future research should aim towards.

## Literature Review

Machine learning (ML) is a subset of artificial intelligence which enables computers, by employing statistical methods, to learn patterns in data and make autonomous decisions [18]. In recent years there has been a great investment in examining the significance of machine learning applicability in predicting chronic diseases, including cardiovascular disease, diabetes, cancer, liver, and neurological disorders [19]. Afsaneh et al (2022) comprehensively reviewed the applications of machine learning in diabetes, disclosing that ML is a promising approach for diagnosis and management of diabetes [20]. Abd et al (2024) assessed ML algorithms, including XGBoost, Random Forest. Decision Tree, Support Vector Machine and Gradient Boosting for diabetic retinopathy (DR), highlighting that ML can facilitate early detection of DR in diabetic patients [21]. Similarly, Ghosh et al., 2025 developed classification models, demonstrating that they can precisely predict a patient’s diabetic status based on various health indicators [22]. Ashisha et al (2023) evaluated five interpretable ML algorithms and revealed the Random Forest technique outperformed all other ML techniques by achieving 98% precision, 98% recall, 98% F1-score, 75% sensitivity, 96% specificity, and accuracy of 97.5% [23]. Metwally et al., (2024) showed that ML can identify sub phenotypes such as muscle or hepatic insulin resistance, β-cell dysfunction, and impaired incretin effect [24], demonstrating the efficacy of ML to uncover underlying features that aid the risk stratification of individuals with early glucose dysregulation. Zargoush et al., (2025) investigated the use of ML for prescriptive analysis, showing that ML can also be used to make personalized treatment decisions [25]. All of this indicates that the real-world applications of these models are increasingly evident and has the potential to benefit clinicians in providing actionable insights, streamlining patient monitoring, and supporting data-driven decision-making.

In addition to finding that ML models are consistently better at predicting diabetes, studies also highlight the importance of feature selection methods [26]. Michelucci, (2024) specifically lists the relevance of feature selection in healthcare regarding hospital readmissions, stating that it helps providers pinpoint the most critical predictors of readmission which can help hospitals on how to optimize management, provide targeted interventions and inform the allocation of resources efficiently [27]. A study by Gürsoy & Alkan, A. (2022) investigated diabetes data with permutation feature importance based deep learning methods and revealed that the HbA1c feature is an important parameter in the model’s classification success [28]. Cahyani & Irsyada, (2025) compared permutation importance vs select best for feature selection across multiple classifiers and found that permutation feature importance led to improved model performance, supporting its use for identifying influential clinical predictors [29]. Della Corte et al., (2025) reports permutation importance alongside SHAP to derive clinically coherent feature reliance patterns in ML models predicting incident diabetes outcomes [30].

Although there has been an evident rise in the use of ML models that have proven effective in enhancing prediction accuracy, their “black box” nature often limits their interpretability and transparency [31]. This lack of explainability makes it difficult for the models to be adopted for clinical use, because healthcare providers must understand the logic behind the decision in order to confidently relay the information to patients. Therefore, to address this limitation interpretable frameworks like SHAP (Shapley additive explanations) have been increasingly incorporated into diabetic prediction studies [32-33]. In large scale population analysis, SHAP has effectively been used to identify key predictors like HbA1c, BMI and age, which demonstrated consistency with what is already established as risk factors in epidemiological studies and reinforced the biological plausibility of ML outputs [34]. Similarly, Prendin et al., (2025) discussed the importance of using tools to interpret the output of black-box models in type I diabetes management and showed that SHAP uncovered patient specific feature dynamics in blood glucose prediction, which could enable clinicians to better understand temporal glucose variability and individualized metabolic responses [35]. Collectively, these findings highlight that SHAP-based explainability improves model interpretability and above all strengthens the bridge between algorithm prediction and clinical reasoning which in turn supports more transparent, reliable as well as patient centered decision support systems in diabetic care.

In this project several models were used: Random Forest, Support Vector Machines (SVMs), Logistic Regression, Decision Tree, XGBoost and Gradient Boosting, all of which are shown to have established effectiveness in medical classification tasks. Random forest is recognized as one of the most powerful and efficient techniques [36]. It was selected because it enables feature importance selection, which supports clinical interpretability by helping clinicians to understand which factors matter [37-38]. Support Vector machines are valuable because they minimize empirical classification errors while increasing geometric margin [39]. Logistic regression is foundational because it is good at understanding the relationship between the dependent variable and independent variables, making it suitable for categorical disease outcomes [40]. Ensemble methods such as XGBoost and Gradient Boosting improve prediction accuracy and generalizability by iteratively improving on difficult-to-classify instances, minimizing wrong predictions [41] and offering hyperparameter tuning [42]. Together these models provide a complementary toolkit for disease prediction and classification.

## Methods

This is a study aimed at predicting diabetes using machine learning into three classes and using explainable artificial intelligence techniques. The data supporting the findings of this study are available online[43]. This study consists of four main steps: First, data was collected and preprocessed to ensure data quality. Next, permutation feature importance was conducted to rank the importance of each feature in diabetes prediction. Then different machine learning models were tuned using RandomizedSearchCV, trained and evaluated to identify the best algorithms for predicting diabetes. Lastly, SHAP technique was employed to interpret the model’s prediction outcomes for 6 cases across the classes and age groups. All the stages of this project are outlined by the graphical abstract shown in **Figure 1**.

**Figure 1.**
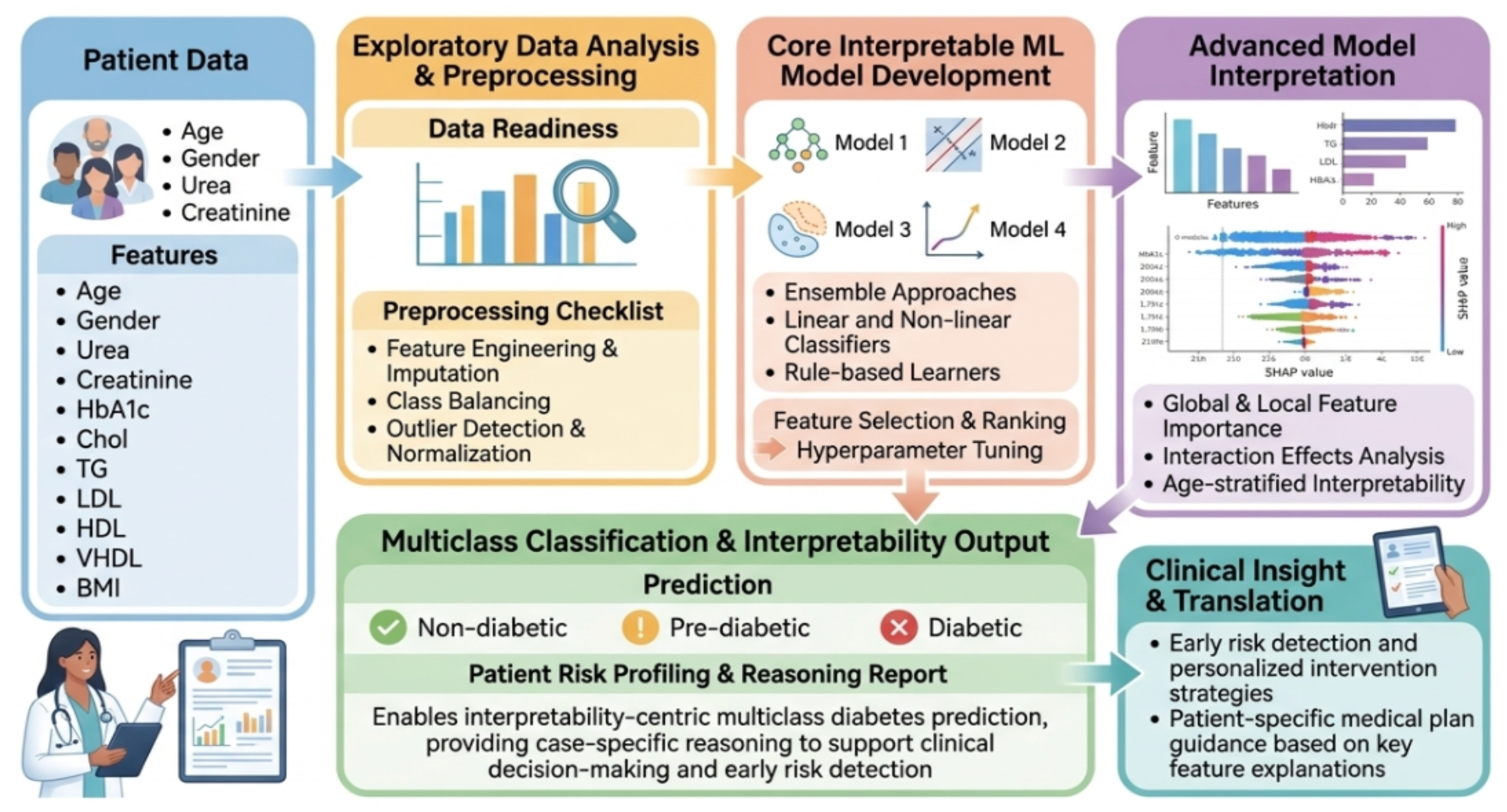
Graphical Abstract. The Explainable ML framework for Diabetic Risk Prediction: Advanced Interpretability-Centric Machine Learning Framework for Multiclass Diabetes Risk Assessment: From Raw Patient Data to Transparent Clinical Action.

### I. Study Population

The data for this study was collected from the Iraqi society by the laboratory of Medical City Hospital and the Center for Endocrinology and Diabetes Teaching Hospital [44]. The cohort comprises health data extracted from patient files into the database constructing the diabetes dataset. Each patient record included demographic variables (age, gender), the anthropometric measure body mass index (BMI) and laboratory measures (haemoglobin A1c, lipid panel, creatinine and urea). A total of 264 individuals were included in the analysis, and these included 129 females and 144 males aged 25 to 77 years. **Figure 2** shows how the categorical variables, gender and diabetes class, were distributed in the dataset.

**Figure 2:**
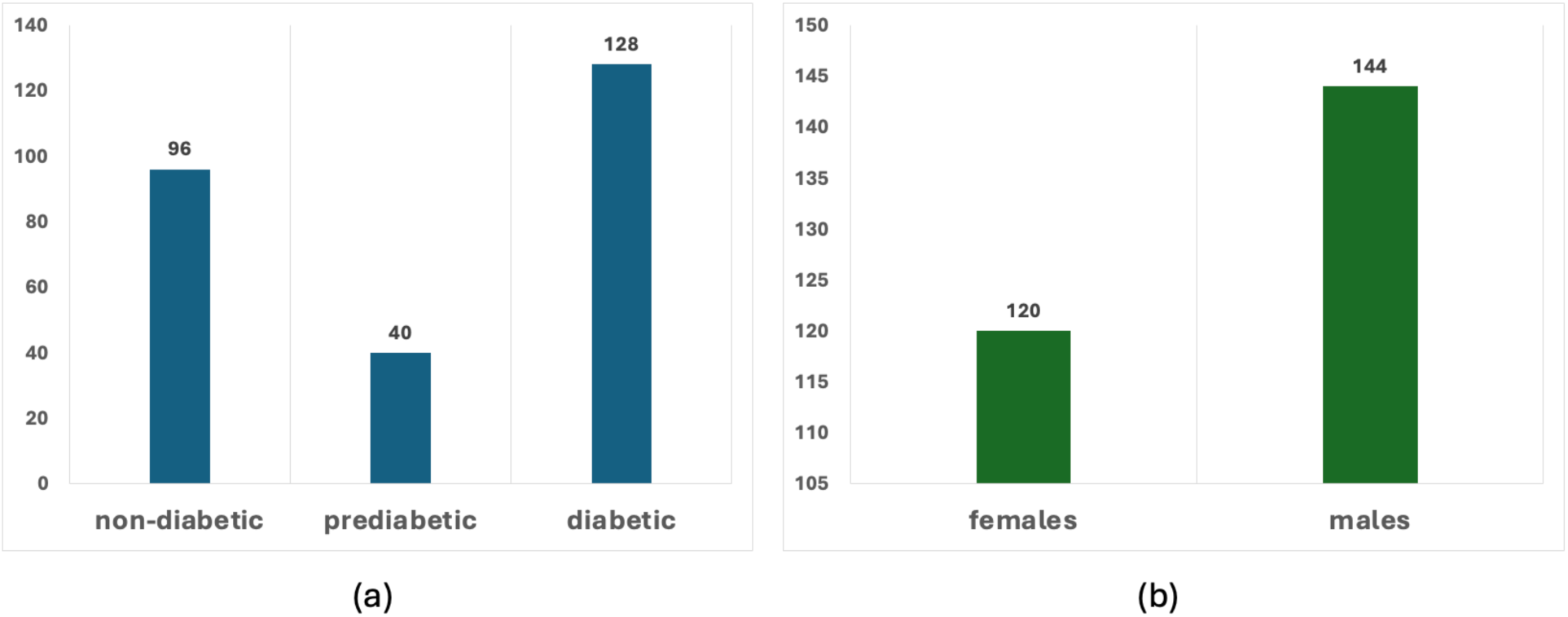
Class and Gender Distribution Analysis. Panel a) presents the distribution of data points across the different diabetes risk classes (non-diabetic, prediabetic, and diabetic). Panel b) displays the distribution of the study cohort by gender (females and males). The numerical values above each bar indicate the specific counts.

### II. Dataset Preprocessing

The dataset contained ten continuous variables (AGE, Urea, Cr’, HbA1c, Chol, TG, HDL, LDL, VLDL, BMI]. Exploratory data analysis (EDA) was done to assess variable distribution. For the continuous variables, a summary statistics table with means, standard deviations, medians and interquartile range were computed. Box plots (**Figure 3**) were created to assess the presence of any outliers and skewness in the data. The box plots revealed that there was a right skewed distribution across a majority of the variables. We see this with variables such as Creatinine (Cr), Urea and VLDL, as demonstrated by the right extending tails of upper outliers. This distribution pattern is typical of medical datasets due to patients experiencing acute or severe conditions. Variables like Age and BMI on the other hand exhibited a tighter, more normally distributed spread. Lastly, continuous variables were standardized in order to make sure that they contributed proportionally to the model training.

**Figure 3.**
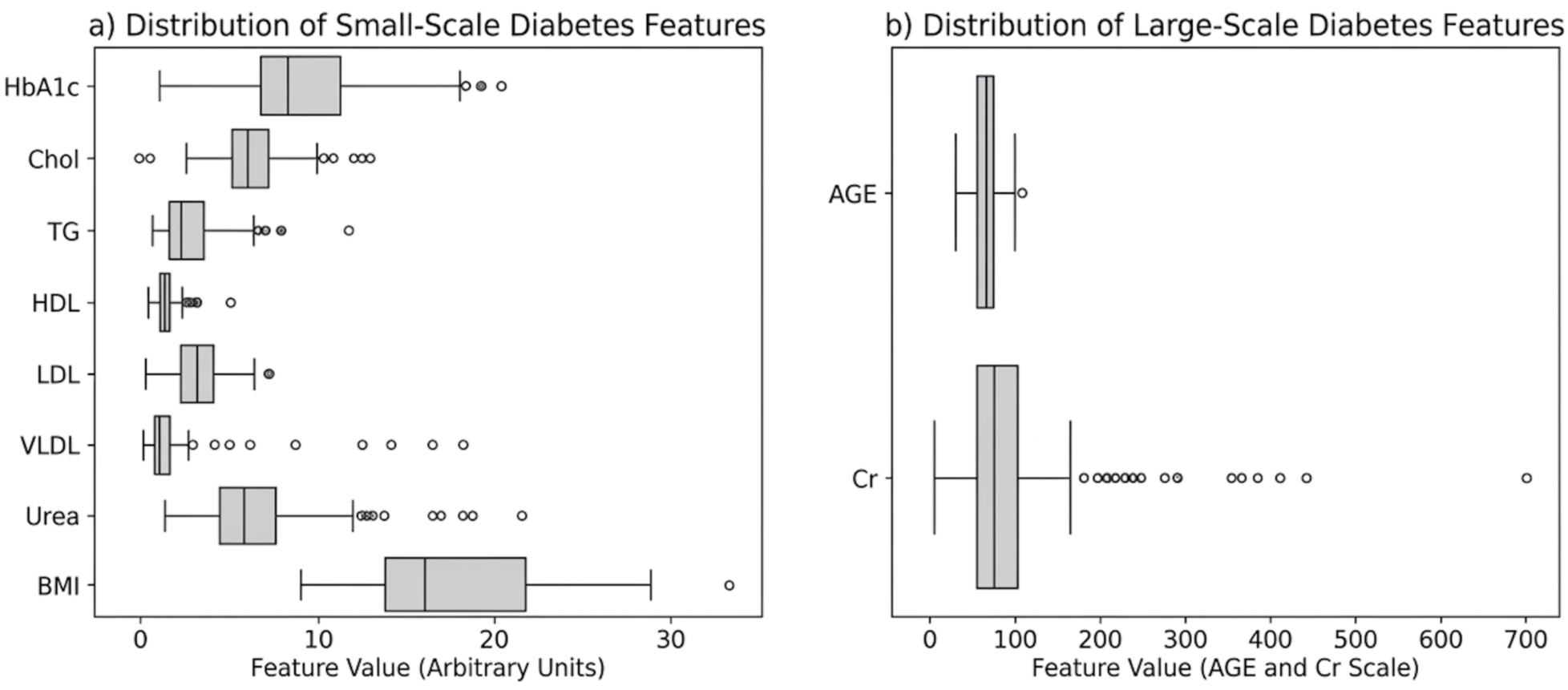
Comprehensive Distribution Analysis of Key Clinical and Biological Features from the Diabetes Dataset. Panel a) (left) displays the distribution of smaller-scale features (HbA1c, Lipids, Urea, BMI), highlighting their variability and outliers. Panel b) (right) shows the wider-scale features of AGE and Creatinine (Cr), demonstrating their distinct, larger value ranges and extreme high-value outliers.

### III. Model Development

A series of supervised machine learning models, Random Forest, Logistic Regression, Support Vector Machine, Gradient Boosting, Decision Tree, and XGBoost, were chosen for classifying patients into 3 diabetes classes. These six models were chosen based on their established performance in medical classification tasks and suitability of tabular health data [45]. Before the models were trained, the dataset was encoded, standardized, split into 80% for training and 20% for testing in order to allow unbiased evaluation of the model’s performance. Proportions of the train and test set were inspected to ensure consistent distribution. The best configurations for each model were found by hyperparameter tuning using RandomizedSearchCV [46]. This was done by predefining the parameters for each classifier, then randomized iterations were executed per model with tenfold cross validation, after which the best hyperparameters identified for each model were re-fit on the full training set before evaluation. **Table 1** summarizes the optimal parameters for each of the utilized models.

**Table 1:** Summary of hyperparameters and optimal values for machine learning classification models.

| Model | Hyperparameter | Tuning Parameters | Optimal Value |
| --- | --- | --- | --- |
| Random Forest | n_estimators: number of trees in the forest | [100, 200, 300] | 200 |
|  | max_depth | [None, 10, 20, 30] | 10 |
|  | min_samples_split | [2, 5, 10] | 5 |
|  | min_samples_leaf | [1, 2, 4] | 1 |
|  | bootstrap | [True, False] | False |
| Logistic Regression | C | [0.1, 1.0, 10.0] | 10.0 |
|  | solver | ['lbfgs'] | lbfgs |
| Support Vector Machine | C | [0.1, 1.0, 10.0] | 10.0 |
|  | kernel | ['linear', 'rbf'] | linear |
|  | gamma | ['scale', 'auto'] | auto |
| Gradient Boosting | n_estimators | [100, 200, 300] | 200 |
|  | learning_rate | [0.01, 0.1, 0.2] | 0.1 |
|  | max_depth | [3, 5, 7] | 5 |
| Decision Tree | max_depth | [None, 10, 20, 30] | 10 |
|  | min_samples_split | [2, 5, 10] | 5 |
|  | min_samples_leaf | [1, 2, 4] | 2 |
| XGBoost | n_estimators | [100, 200, 300] | 300 |
|  | learning_rate | [0.01, 0.1, 0.2] | 0.01 |
|  | max_depth | [3, 5, 7] | 7 |
|  | colsample_bytree | [0.7, 0.8, 1.0] | 0.7 |
|  | subsample | [0.7, 0.8, 1.0] | 0.7 |

### IV. Evaluation Metrics

Model performance was quantified using the following metrics. For the equations TP, TN, FP, and FN denote true positives, true negatives, false positives, and false negatives, respectively.

Accuracy: Measures the amount of correct predictions the model made. It is defined as:

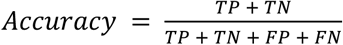

Precision: Gives the ratio of true positive predictions to the total predicted positives. Its equation is:

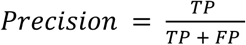

Recall: Reflects the ability of the model to identify all relevant instances. It is calculated as follows:

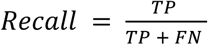

F1-score: Balances the precision and recall metrics giving us the harmonic mean of both these metrics. It is very useful in imbalanced data sets as it balances the tradeoff between false positives and false negatives. It is calculated as:

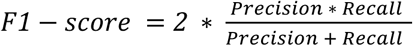

For the multiclass setting, these metrics were computed using macro-averaging so that each diabetes class contributes equally to the final evaluation, regardless of the respective class sample size.

### V. Feature Importance

The project also conducted a feature importance analysis to rank features based on how well they predicted outcomes [47]. Feature importance was evaluated using both Random Forest Mean Decrease in Impurity (MDI) and permutation importance. The MDI measures the contribution of each feature to impurity reduction during tree construction and permutation importance assesses the decrease in model performance when feature values are randomly shuffled, providing a model-agnostic estimate of feature relevance. This makes it easy to comprehend how the key characteristics of diabetes dominantly impact the model’s performance.

### VI. Demographic Stratification

Subgroup analyses by age and gender were also conducted to evaluate the model’s performance across clinically relevant subgroups. For age, two age strata were formed using the recommended diabetic screening cut-point of 35 years [48-49]. We then categorize gender into male and female. The strata were applied to the entire dataset prior to the model being trained. A Gradient Boosting classifier was trained and evaluated following the same pipeline implemented in the primary analysis.

### VII. Explainable Artificial Intelligence (XAI) Techniques

The SHAP method was used to help the interpretability and transparency of the models. SHAP uses Shapely values to explain the contribution of each feature on the model’s prediction. A Shapely value is described as the average marginal contribution of a feature value across all possible coalitions [50]. Knowing how much each feature contributes to the model’s decision allows clinicians to understand what drove the prediction for that individual. SHAP was chosen because it is good at horning in on specific model behaviors with more precision, which helps boost trust and interpretability. Kabir et al., (2025) discusses how there exists a tradeoff dilemma between accuracy and interpretability with highly accurate deep learning models due to their complexity which often leads users to accept reduced performance in exchange for more interpretable models, however techniques like SHAP help balance this trade off [51]. Similarity, Paliwal et al., (2025) emphasizes that explainable AI techniques bridge the gap between model complexity and user trust because they highlight detailed, targeted explanations on these high performing models [52]. SHAP was chosen over other explainable techniques because it has a strong theoretical foundation rooted in Shapley values, ensuring fair distribution of feature contributions [53]. Moreover, SHAP can be used on any supervised ML model and is more robust and consistent compared to LIME which bases its explanation on random sampling often leading to multiple explanations for the same instance [54]. Additionally, SHAP offers both local and global interpretability, which means they don’t only provide the interpretability for one prediction but also how the model functions as a whole [55]. These advantages support what has led to the widespread utilization of SHAP in literature especially in key domains like healthcare.

## Results

Six models were trained, tuned and evaluated to classify diabetes status. Across all evaluated classifiers, ensemble-based methods achieved the strongest predictive performance for multiclass diabetes classification. Results are visualised in **Figure 4**. XGBoost emerged as the top-performing model, achieving the highest accuracy (1.00), precision (1.00), recall (1.00), and F1-score (1.00), indicating near-perfect performance on this dataset, likely influenced by dataset size and structure. The Random Forest model also performed strongly, with an accuracy of 0.962, recall of 0.917 and an F1-score of 0.940. Gradient Boosting demonstrated moderate performance, achieving an accuracy of 0.906 with balanced precision (0.912) and recall (0.893), suggesting stable classification capability. In contrast, the Support Vector Machine and Logistic Regression models showed comparatively lower performance, particularly in precision and F1-score, reflecting reduced effectiveness in handling class boundaries within the multiclass setting. The Decision Tree model performed better than linear models but was inferior to ensemble approaches, highlighting the advantage of ensemble learning methods in capturing complex nonlinear relationships inherent in diabetes progression.

**Figure 4.**
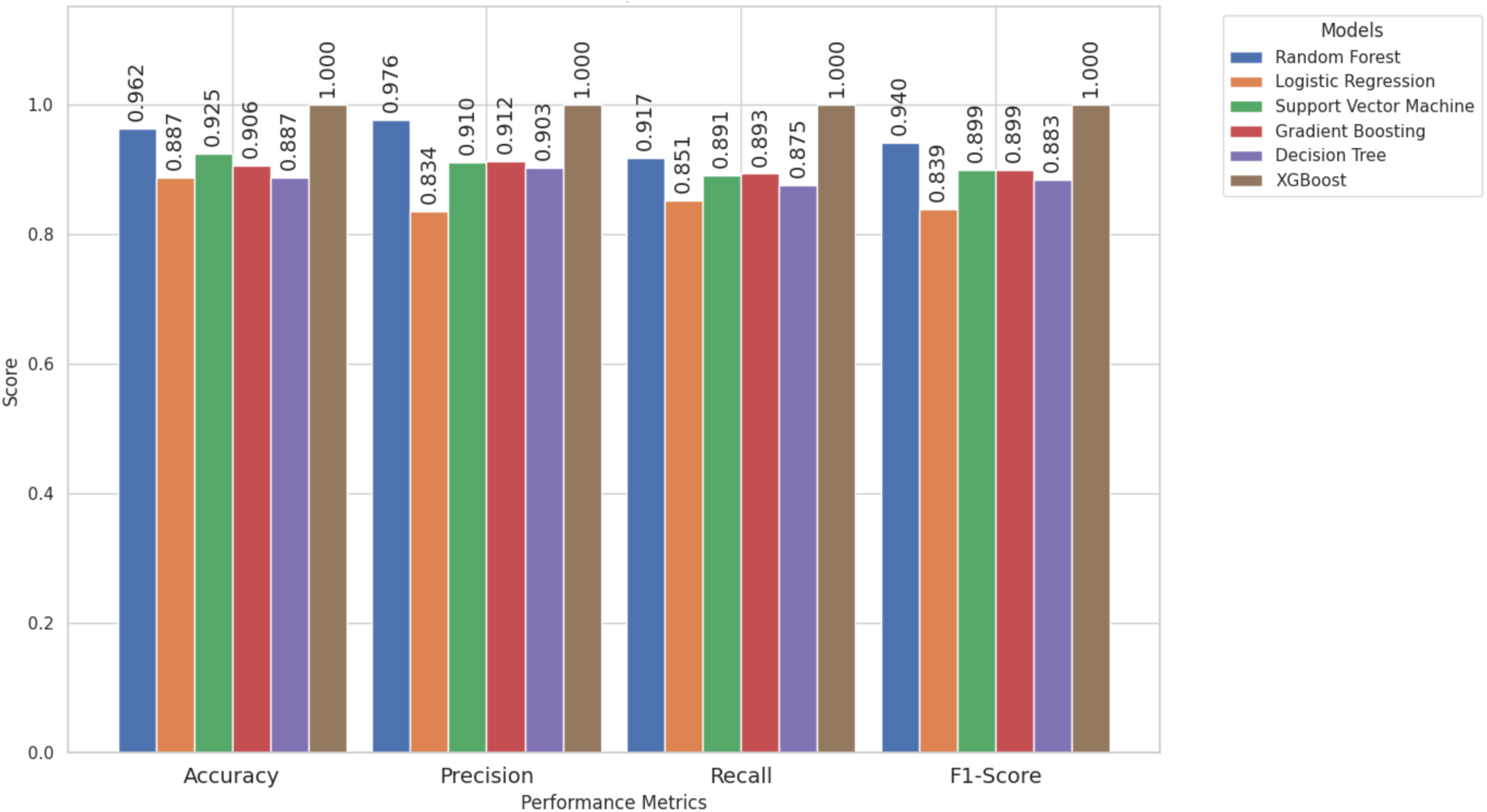
Comparative evaluation of machine learning model performance across key classification metrics (Accuracy, Precision, Recall, F1-Score). The bar chart provides a side-by side evaluation of the models. Numerical annotations above the bar provide exact performance coefficients, facilitating a direct assessment of model reliability and classification precision.

To further evaluate how well the model generalizes to unseen data, we compared the training, cross-validation (CV), and test accuracies across all evaluated models (**Figure 5**). Gradient Boosting emerged as the most robust on unseen data with a test accuracy of 0.981, training score of 0.991 and cross validation score of 0.972. The Gradient Boosting remained stable across the three phases, showing great generalization capabilities without overfitting compared to the XGBoost. The XGBoost model achieved the highest training accuracy of 1.00, a cross-validation score of 0.986 However, the test score accuracy decreased to 0.962, indicating slight overfitting. The Random Forest and the Decision Tree had moderate strong performance, displaying similar generalizability patterns. The Random Forest started with a slightly higher training score than the decision tree (0.981 vs 0.972) but both converged to the same validation score of 0.962 and final test score of 0.962. The SVM had the sharpest drop in performance as it started with a strong training score of 0.972, resulting in the lowest test accuracy of 0.865. Logistic Regression on the other hand still yielded consistently lower metrics across the three phases, starting at 0.948 in training and finishing at 0.885 on the test set. Support Vector Machine (SVM) and Logistic Regression struggled the most with generalization. Overall, ensemble-based methods demonstrated superior predictive performance and generalization compared to single linear models.

**Figure 5.**
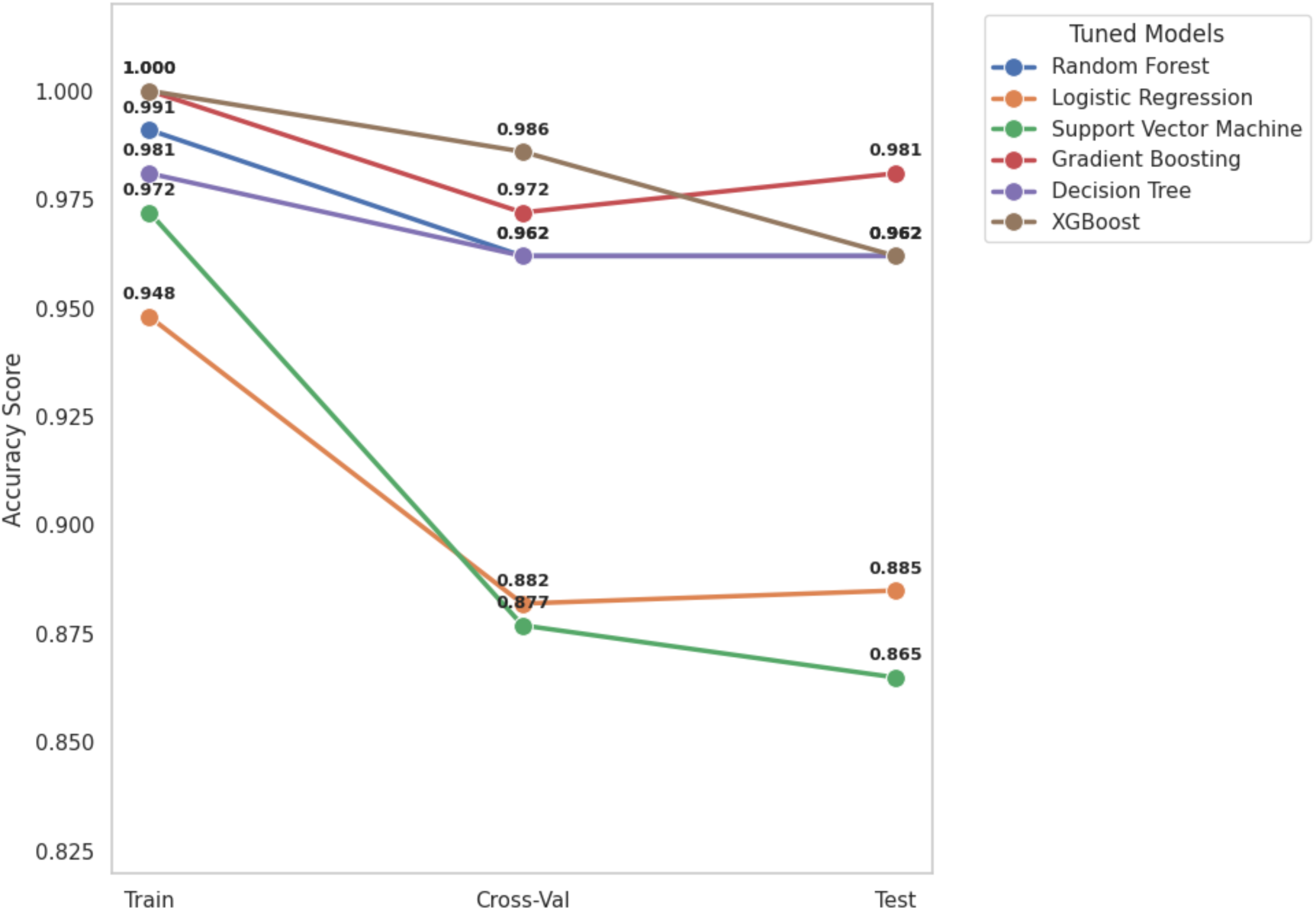
A variational accuracy scores of tuned classification models across train, cross-validation, and test diabetes datasets. The line graph illustrates the performance trajectory of the optimized models (each corresponding to a specific color) from training through cross-validation to final testing. High performing models like Gradient Boosting maintained superior accuracy levels in the test dataset (0.981) indicating strong generalization capabilities. In contrast, Logistic Regression and Support Vector Machine exhibit a more pronounced performance decline following the training phase. Individual data point labels provide precise accuracy coefficients for each validation stage.

The Gradient Boosting, which emerged as the top robust model, was used to construct ROC curves and results are shown in **Figure 6**. The AUC-ROC represents the model’s ability to distinguish between classes across various thresholds. When the AUC value is high it indicates that the model is performing better. Due to the small test set sample size, averaging ROC curves across multiple Stratified Fold splits was done because it aggregates information from all the folds rather than relying on a single limited test set, hence producing a more reliable ROC curve. The results show outstanding classification performance with slight variations between the classes. Class 0 (non-diabetic) had the highest AUC of 0.991, followed by Class 1 (prediabetic) with an AUC of 0.986 and lastly Class 2 (diabetic) with an AUC of 0.972. The diabetic class was comparatively the challenging class for the model to classify as shown by the green solid line sitting lower than the others. Overall, the model was highly effective at distinguishing between the three classes, with all the AUC scores above 0.97.

**Figure 6.**
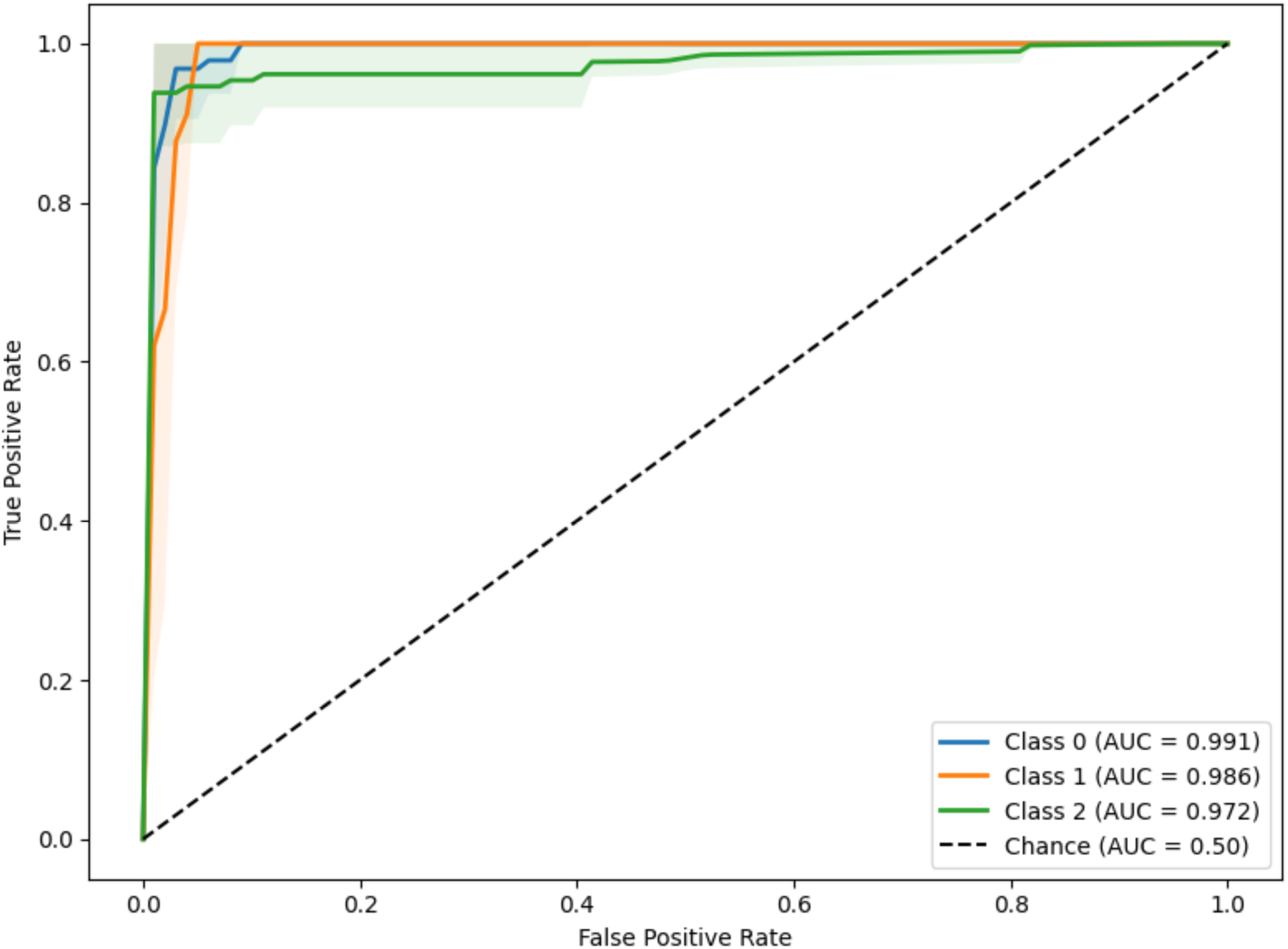
Receiver Operating Characteristic (ROC) Analysis for the tuned Gradient Boosting Classifier. The plot displays the cross-validated ROC curves for the three diabetes classes risk categories. Class 0 (non-diabetic), Class 1 (prediabetic), and Class 2 (diabetic). High Area Under the Curve (AUC) values ranging from 0.972 to 0.991, demonstrate the model’s powerful discriminatory power in distinguishing between classes. Shaded regions surrounding each curve represent the variance across validation folds, dashed diagonal line indicates the baseline for random chance (AUC = 0.50).

The feature importance analysis showed that the model’s decision-making process is driven by three core clinical indicators, and all the other variables contribute very little. The key predictors are HbA1c, BMI and Age. HbA1c exhibited the highest MDI (exceeding 0.5) and Permutation importance (approximately 0.45) score, indicating that the model relies fundamentally on the patient’s HbA1c level to make a classification. The permutation score indicates that removing or shuffling this feature would severely degrade the model’s accuracy. The analysis also ran the models without the HbA1c to assess how it impacts the accuracy of the metrics and there was an observed decline in the accuracy across all classifiers, with Random Forest decreasing from 0.962 to 0.906, Logistic Regression from 0.887 to 0.774, Support Vector Machine from 0.926 to 0.717, Gradient Boosting from 0.906 to 0.868, Decision Tree from 0.887 to 0.830, and XGBoost from 1.00 to 0.887 as shown in **Figure. 8**. These findings underscore the dominant role of HbA1c as a predictive biomarker and highlight its critical contribution to model performance in diabetes classification. BMI and Age were the only other features that offer meaningful predictive value. BMI was the second important feature with moderate influence (MDI ∼ 0.18, Permutation ∼ 0.08). Age ranked third, contributing a smaller but recognizable importance to the model (MDI ∼ 0.09, Permutation ∼ 0.03). This is supported by the correlation heatmap in **Figure 9**, whereby HbA1c and class had the strongest positive correlation of 0.77, followed by BMI and class with a correlation of 0.75 then age and class with a positive correlation of 0.51. The remaining eight features (Urea, Chol, VLDL, Cr, TG, Gender, LDL, and HDL) showed minimal to no predictive power, all with MDI scores below 0.05 and permutation scores at zero (indicated by lack of the orange bars).

**Figure 7.**
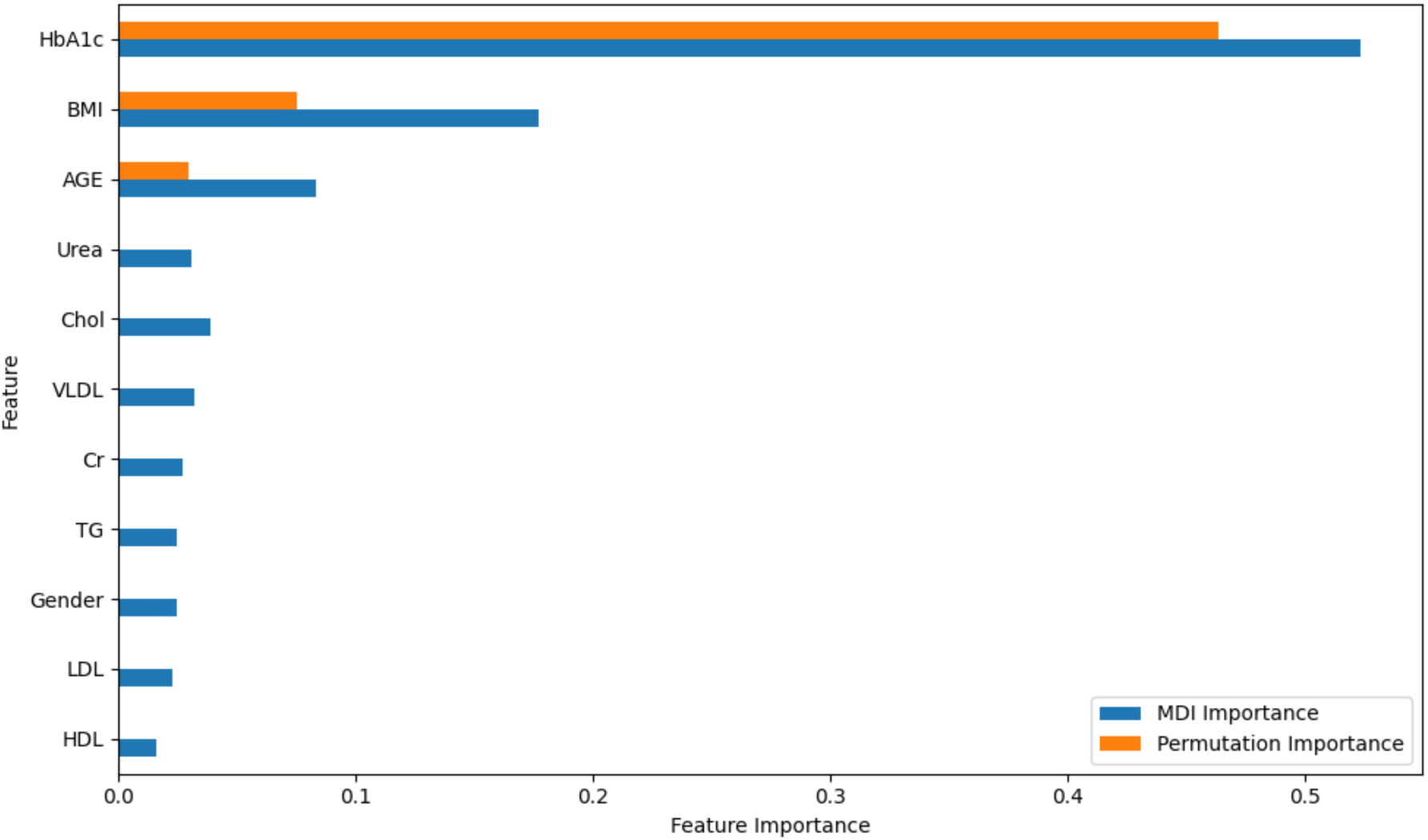
Feature Importance ranking and Mean Decrease Impurity (MDI) Analysis. This horizontal bar char evaluates the relative contribution of clinical and biological variables to the machine learning model’s predictive performance. HbA1c emerges as the primary predictor by a significant margin in both MDI and Permutation Importance scores, followed by BMI and AGE. The comparison reveals that most other biological features link Urea, Cholesterol and Lipid biomarkers have lower importance, signifying that the model’s sensitivity lies heavily on glycemic control and demographic factors like AGE.

**Figure 8.**
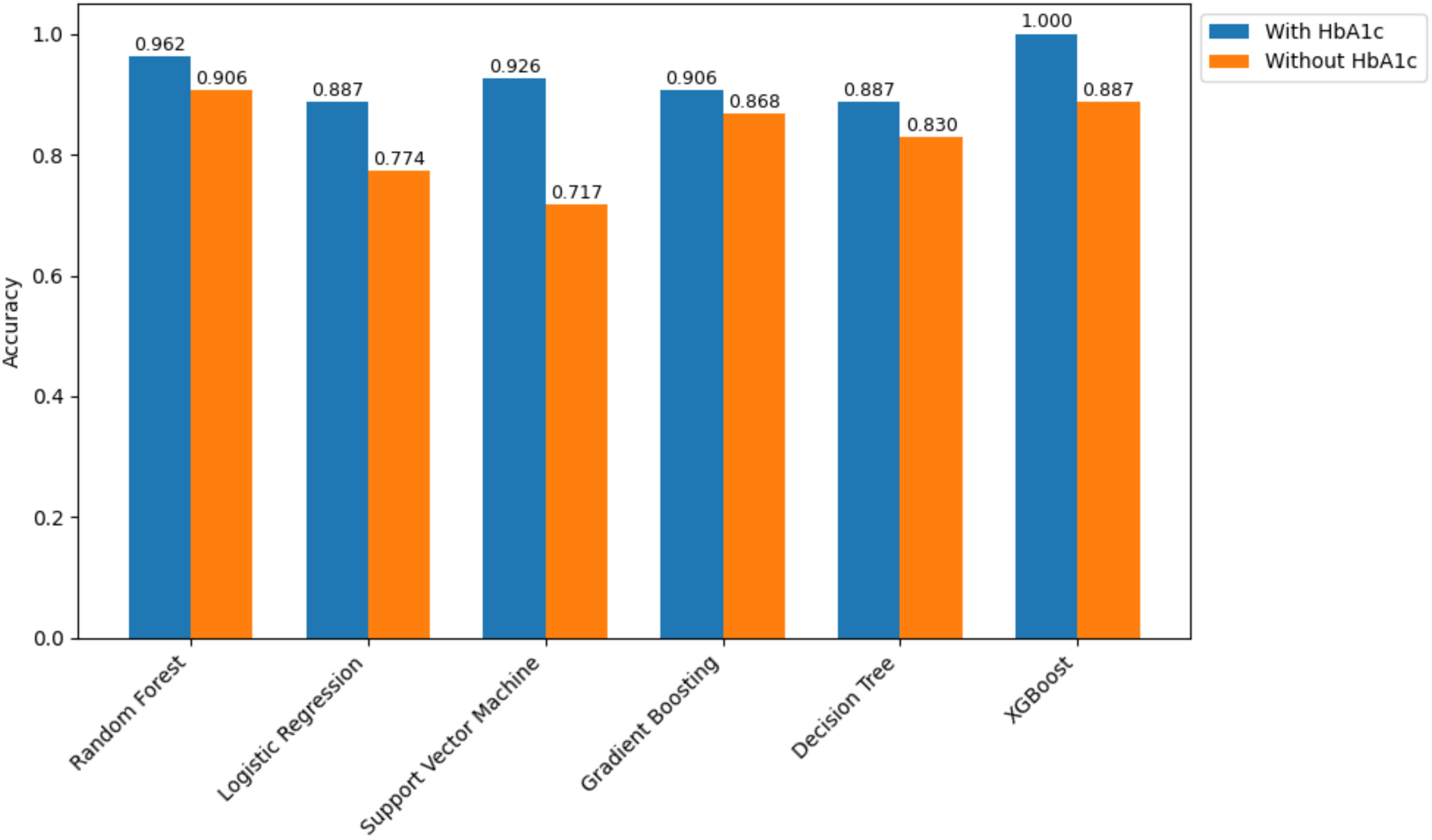
Sensitivity Analysis of HbA1c as a critical predictive feature on Model Performance. The grouped bar chart illustrates the differential impact on model accuracy when the HbA1c feature is removed from the dataset. A consistent performance decline across all six models, most notably in the Support Vector Machine and Logistic Regression. Where accuracy drops by approximately 0.209 and 0.113, respectively. The sustained accuracy of the XGBoost and Random Forest Models (above 0.88) even after feature removal highlight their robustness.

**Figure 9.**
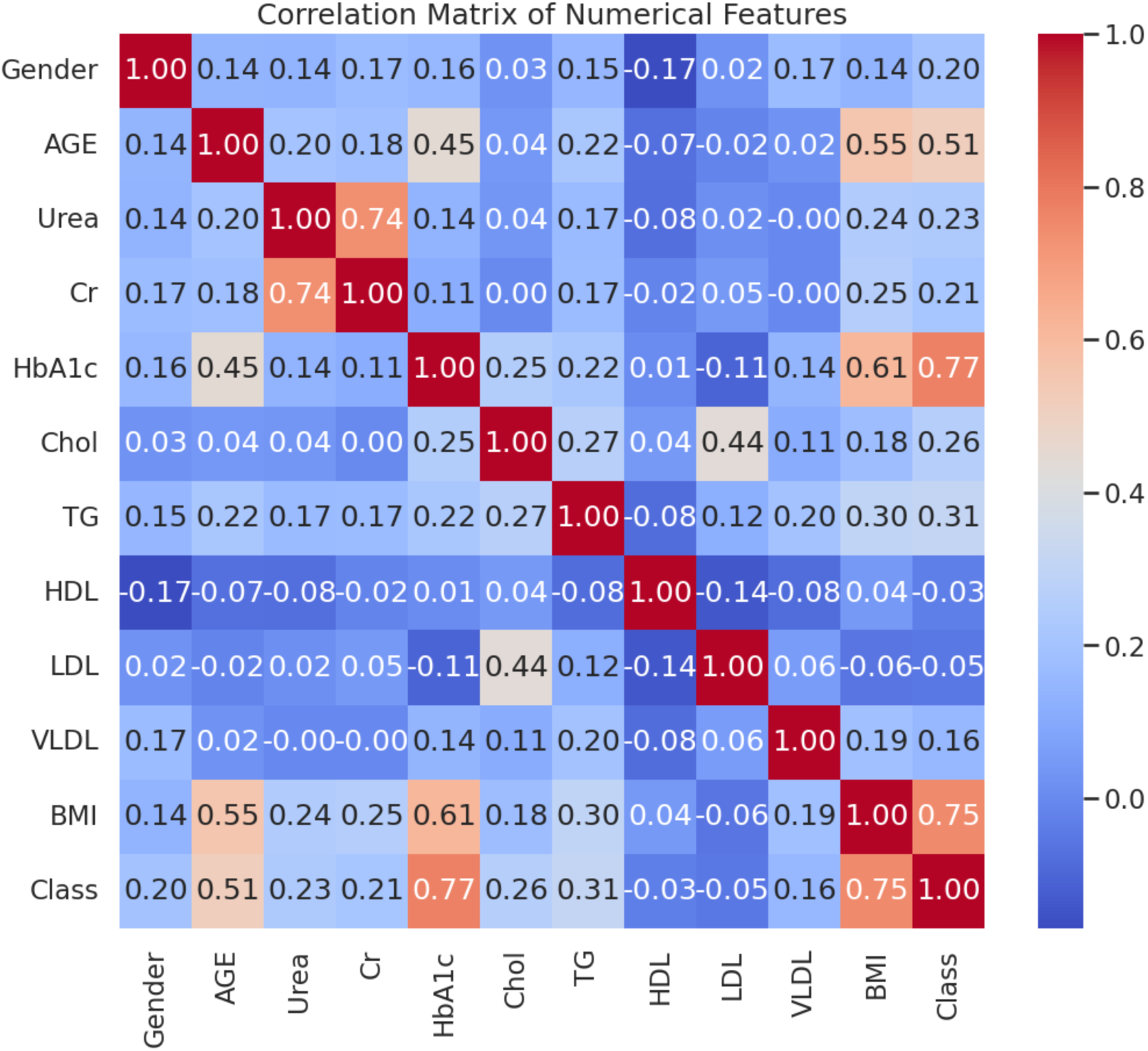
Pearson Correlation Heatmap of Clinical and Biological Features. This matrix visualizes the strength and direction of linear relationships between all numerical variables in the study cohort. Armstrong positive is observed between the Class variable and HbA1c (0.77), BMI (0.75), confirming their roles as significant predictors. Color ranges from blue (negative correlation) to red (positive correlation).

To evaluate the model for potential demographic bias, the performance was analyzed across age and gender. Though the analysis included gender, it is important to note that the focus was placed on age because the feature importance analysis revealed gender to be negligible on the model’s performance. The subgroup breakdown by age revealed some performance disparities. **Figure 10** shows that the model performed consistently well on older individuals, achieving an accuracy of 0.94, F1 score of 0.92, which is slightly higher than the overall metrics on the whole dataset’s test set. Performance on younger individuals was skewed as it achieved a perfect accuracy of 1.00, however the precision, recall and F1 score dropped to 0.67. This imbalance is due to the severe class imbalance and extremely small sample size (n=5), hence it would make it difficult for the model to reliably generalize for younger patients. These findings suggest that stratified models provide more reliable predictions in well-represented populations, partially supporting the hypothesis that demographic stratification improves model performance.

**Figure 10.**
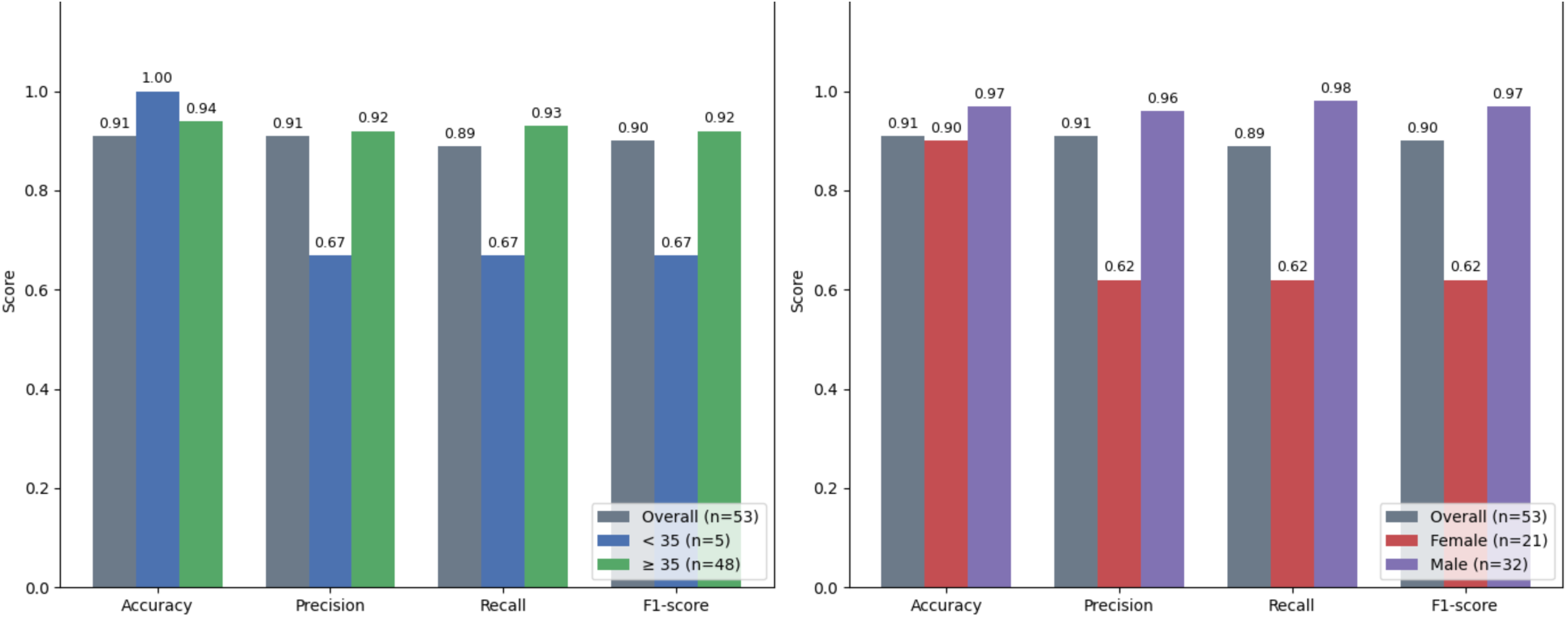
Comparative Summary of Stratified Model Performance Metrics across age and gender subgroups vs on the Overall dataset. The bar chart provides side by side analysis of performance predictors (Accuracy, Precision, Recall and F1-Score) for age strata vs the whole data (on the left) and the gender strata vs the whole dataset (on the right). This shows how effectively the model maintains classification across different subsets of the dataset, in order to determine whether the model’s efficacy is uniformly distributed or specific to stratified characteristics.

SHAP analysis was used to interpret the multiclass prediction model in order to identify key clinical factors of the model output across the classes (non-diabetic, prediabetic, diabetic). The SHAP summary plot (**Figure 11**) shows the overall impact of each feature on predicting the class. HbA1c stood out as the strongest driver for diabetes class classification although the magnitude and direction of influence varied for each diabetes stage.

**Figure 11.**
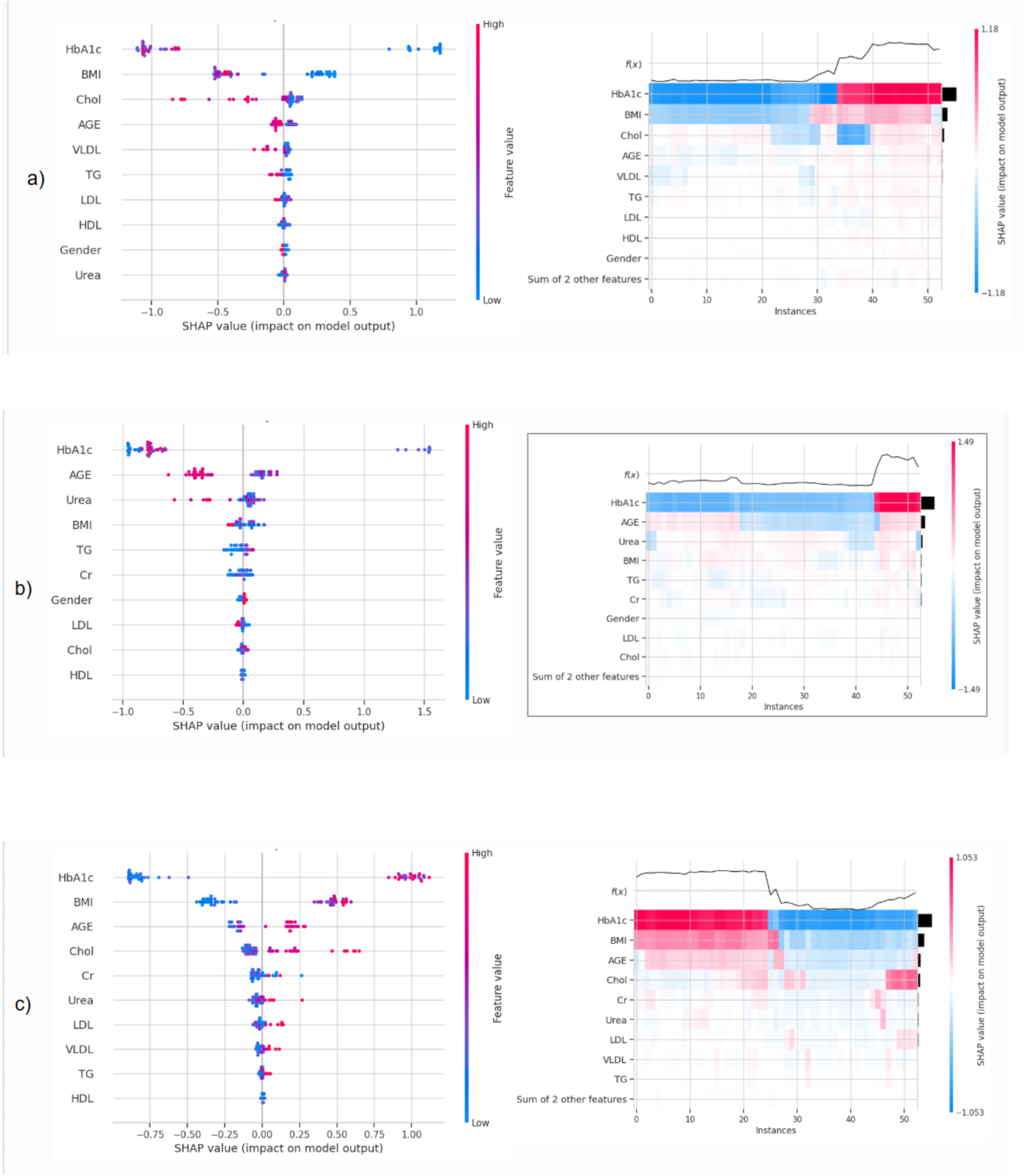
Class-specific SHAP Summary Plots and Feature Contribution Heatmaps for Diabetes Classification. The visualization breaks down the model’s decision-making process into three panels representing (a) non-diabetic, (b) pre-diabetic, and (c) diabetic classes. Features are ranked by their predictive importance, with color gradients illustrating the relationship between high and low feature values and their corresponding positive or negative influence on class prediction. This reveals how the diagnostic significance of specific features like HbA1c shift across different classes of diabetes, offering a more interpretable view of the model’s classification logic.

For the non-diabetic class, summary plots showed that lower levels of HbA1c were strongly associated with positive SHAP values, indicating that lower values pushed the prediction towards the non-diabetic class. Additionally, the plots for the non-diabetic class demonstrated moderate influence with BMI and cholesterol, here higher BMI pushed the prediction away from being non-diabetic. The corresponding heatmaps supported these findings, HbA1c maintained consistent dominant influence across individuals, providing robustness of feature importance.

In the prediabetic class, HbA1c as well as age showed the strongest and most variable SHAP distributions as high values seemed to have bidirectional influence on the prediction, which is expected given the transitional state of the disease at this stage. Age and BMI also had bidirectional contributions to the model’s classification. The heatmap confirmed that these features sustained influence across most individual instances even though their impact was more varied than in the non-diabetic population.

Lastly, for the diabetic class, high values of HbA1c produced strong positive contributions to diabetic predictions. BMI and age also showed strong positive contributions to prediction outcomes. Additionally in this class, lipid biomarkers such as Cholesterol also showed some contributions to the prediction, with higher levels pushing the prediction to diabetes class.

Overall, the SHAP summary and heatmap analysis reveal that, especially for HbA1c and BMI, there are strong feature-specific turning points that could be translated into clinically interpretable insights on how individual risk factors drive diabetes classification across different stages. This supports the potential use for personalised risk stratification and early intervention.

Although **Figure 11** gives us a global view of what features matter across the whole population, **Figure 12** takes a step further and shows how the features play out for six real individuals. Six representative patients were selected across the three disease classes and age groups for SHAP force plot analysis. These visualizations show which specific clinical features contribute to the final classification in relation to the model’s base value. In each case, features shown in red push the prediction toward the predicted class, while features in blue push the prediction away from it. The final model output f(x) reflects the combined effect of all feature contributions, with higher values indicating stronger support for the predicted class.

**Figure 12.**
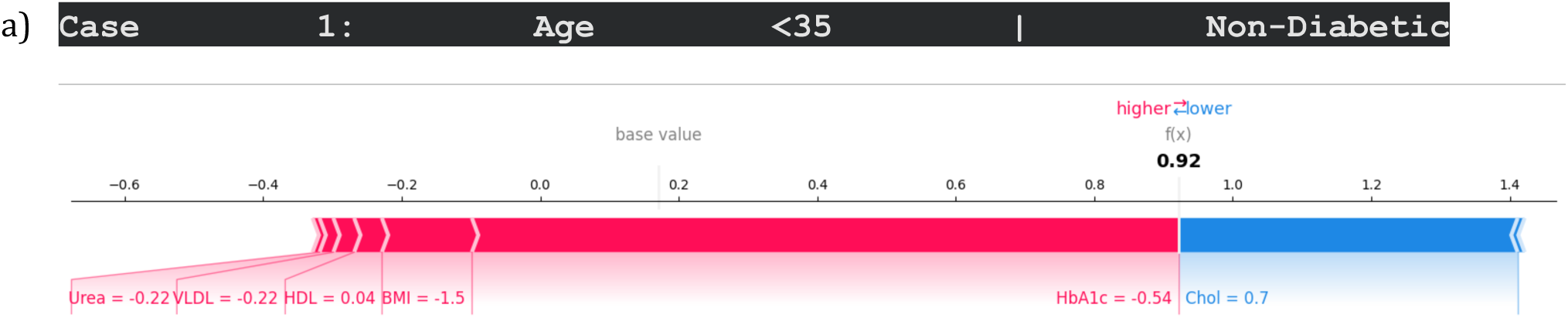
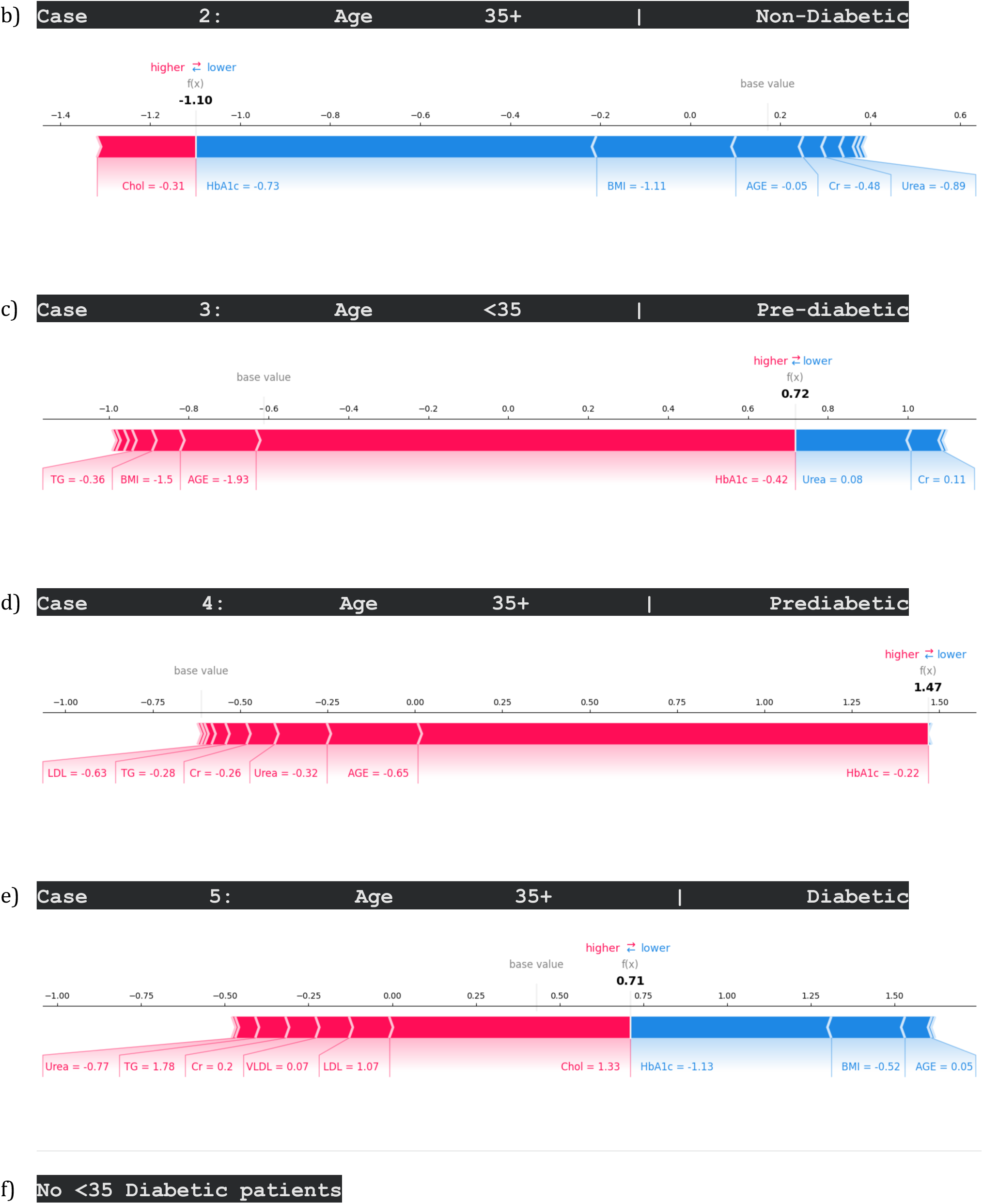
SHAP Force Plots for Individual Patients Across Individual Patient Cases and Age Groups. Panels (a-f) provide a granular visualization of how specific clinical features “push” the model’s prediction away from or towards a specific class output for representative individuals.

### Case 1: Age <35 | Non-Diabetic

The model shows high confidence in making this prediction, with a final output f(x) = 0.92. The prediction was pushed towards the non-diabetic class by primarily lower BMI and lower HbA1c. A higher Cholesterol, on the other hand, was the only feature that pulled the prediction lower but it was heavily outweighed by the protective factors (BMI and HbA1c). This case depicts that glycemic control and body composition have a dominant effect in determining this individual’s low risk to diabetes.

### Case 2: Age 35+ | Non-Diabetic

Contrary to the younger patient, this patient had a model output f (x) = 1.10, which indicates that the model had lower confidence in classifying case 2 into the non-diabetic class. For this case even though the patient was non-diabetic, they were still flagged for having low renal markers like urea and creatinine. A lower cholesterol acted as a protective factor from diabetes prediction. This case suggests that, in older individuals there are additional clinical biomarkers beyond glycemic control that influence the model’s prediction.

### Case 3 : Age <35 | Pre-Diabetic

The model predicts prediabetes with a moderate confidence f(x) = 0.72. The key contributing factors that pushed the prediction to the prediabetic class were a lower-than-average HbA1c, Age, BMI and Triglycerides. However, elevated urea and creatinine slightly pushed the prediction away from the prediabetes stage. This case reveals that prediabetes is characterized by more variable feature interactions.

### Case 4: Age 35+ | Pre-Diabetic

The prediction is driven primarily by a lower-than-average HbA1c, Age, urea. Additionally, triglycerides and LDL pushed the prediction to the prediabetic class. The combination of these features reflects early metabolic dysregulation, suggesting that there are higher risk patterns in the older individual compared to the younger prediabetic case.

### Case 5: Age 35+ | Diabetic

The model confidently classified the individual as diabetic f(x) = 0.71. The factors that drove the predictions to the diabetes class were elevated cholesterol and higher LDL levels, even though their HbA1c was lower than average. Non-regulated triglycerides, urea and creatinine additionally acted as indicators for diabetes for this case. This aligns with clinical patterns where dyslipidemia is seen as a major indicator of diabetes progression.

### Case 6: Age <35 | Diabetic

There were no individuals under 35 years in the dataset that were classified as diabetic, which limited the ability to evaluate SHAP-based explanations for this subgroup. This further highlights the impact of class imbalance and sample size limitations on subgroup interpretability.

The force plot analysis supports the hypothesis that stratified models are more valuable in model interpretability. The exact same types of values (lower BMI and HbA1c) have opposite impacts based on the patient’s age and overall profile (evidenced by case 1 vs case 2), proving that a "one-size-fits-all" threshold is less effective than a personalized SHAP-based assessment. These findings demonstrate that diabetes risk is caused by complex, non-linear interactions rather than by fixed thresholds, supporting the need to use stratified, explainable ML approaches in order to improve personalized risk assessment.

## Discussion

This study evaluates whether demographic stratification improves multiclass diabetes classification compared to universal model approaches. First, the study discovered that ensemble methods, particularly Gradient Boosting, XGBoost and Random Forest, are better suited to modeling the non-linear interactions driving diabetes progression across the three stages: non- diabetic, prediabetic and diabetic. Prior literature supports these model’s superior performance, with studies such as Gangani et al. (2025) showing that XGboost and Random Forest consistently achieve higher predictive accuracy, outperforming linear and single tree models, especially when paired with effective feature selection strategies [56]. The strong performance exhibited by the XGBoost highlights the importance of employing ensemble techniques for multiclass diabetes prediction [57], especially for metabolic diseases like diabetes that have overlapping clinical profiles like prediabetes [58-59]. Contrary to this, weaker performing models like Support Vector Machines and Logistic Regression, are known to be limited in capturing the non-linear boundaries between diabetes disease stages [60], all of which re-enforce the need for more flexible modelling frameworks in clinical risk stratification.

Age based stratification incited us into model reliability and interpretability. The Gradient Boosting model showed robust performance in the 35+ age group, supported by a larger sample size and complete class representation. The slight reduction in recall for the diabetic class suggests that some advanced cases may still be misclassified, potentially due to overlapping metabolic profiles between late-stage prediabetes and early diabetes [61-62]. In contrast, although perfect accuracy was observed in the <35 age group, the very small sample size and absence of diabetic cases severely limited the interpretability and reliability of these findings. This discrepancy emphasizes that stratification can improve interpretability and contextual relevance, but only when supported by sufficient and representative data [63].

SHAP-based model interpretation further reinforced the clinical plausibility of the learned patterns [64-65]. Across all classes, HbA1c, BMI and age emerged as dominant predictors, consistent with established clinical knowledge [66-68]. Importantly, the direction and magnitude of these features varied by disease stage, reflecting the progressive nature of diabetes. For example, HbA1c exhibited clear stage-specific turning points, with lower values demonstrating protective effects in the non-diabetic class and higher values demonstrating the opposite in the diabetic class. The bidirectional behavior of age and BMI in the prediabetic class highlights the heterogeneity of this intermediate stage, where metabolic dysregulation may precede overt glycemic deterioration [69].

Force plot analysis provided further evidence that stratified, patient-level explanations offer meaningful clinical insights beyond aggregate metrics. Individual predictions revealed distinct metabolic risk phenotypes across age groups and disease stages, demonstrating that similar outcomes can arise through different feature pathways. These findings directly support the study’s hypothesis that stratified machine learning approaches enhance interpretability and personalized risk assessment compared to a single universal model.

Several limitations should be addressed pertaining to this study. First, the dataset had low representation of individuals under 35 years, resulting in decreased generalizability of the stratified analysis for these underrepresented subgroups. Although oversampling algorithms like SMOTE were considered to address class imbalance and small subgroup sizes, they were not used for this model pipeline to preserve real-world applicability and avoid artificially inflating predictive performance. Since the goal of this project is clinical translation, reliance on synthesized samples limits external validity [70-72]. Second, while SHAP values improve model interpretability, they only reflect the model’s learnt associations, not causal correlations. Thirdly, while this study used clinical and laboratory variables, the progression of diabetes is also impacted by lifestyle factors like diet and physical exercise. Future studies should incorporate these lifestyle factors into predictive models to enhance predictive depth. Expanding the dataset, integrating longitudinal follow-up data and doing external validation on an independent dataset would test the model’s reproducibility and further support clinical adoption. Additionally, integration with electronic health records systems and clinical trials would help in determining the practical implementation for early diabetes prediction and patient outcomes in a real-world clinical setting.

### Techniques Public Health Implications

Currently clinical screening approaches rely on universal diagnostic threshold, which often fail in capturing individual variability across demographic subgroups such as age and sex [8,49]. From a public health lens, this limitation reduces the effectiveness of early detection strategies as it may result in patients being misdiagnosed further causing unnecessary treatment or delayed diagnosis. To address this issue, it is very important to consider the diverse risk factors and how they would vary in different populations when developing screening programs [73]. This study proposed and found that stratified models produced interpretable results and distinguished between risk factors that vary from individual to individual.

Despite the demonstrated strong predictive performance of ML models, there is still an increasing need for clinically interpretable and transparent decision-support tools that can be adopted into healthcare’s routine practices [31]. This study therefore proposes a stratified and explainable machine learning framework for diabetes classification. The integration of SHAP-based explanations enables transparent identification of key risk factors, such as HbA1c, BMI, and age, at both the population and individual levels. Hence, by presenting these sub-group specific metabolic patterns, clinicians would be able to provide accurate predictions with clear patient-specific explanations to patients when they inquire about what mainly made them susceptible to diabetes.

Moreover, although ML approaches support earlier and more precise risk assessment, there is still a limited translation into healthcare workflow. Hence, integration such a model pipeline into an electronic health record system could allow for proactive risk identification, targeted preventive counselling and earlier referral to lifestyle or metabolic intervention programs. Overall, this study contributes to the development of equitable, data-driven public health strategies by demonstrating that demographic-aware and explainable machine learning models can enhance early detection, support personalized care, and ultimately improve population-level diabetes outcomes.

## Conclusion

This study demonstrates that stratifying machine learning models by demographic factors, particularly age, revealed heterogenous feature influence patterns, supporting the hypothesis that demographic aware model frameworks enhance clinical interpretability. Ensemble models such as Gradient Boosting, XGBoost and Random Forest achieved high predictive performance, while explainable AI techniques revealed biologically and clinically meaningful feature contributions across disease stages. The results show that diabetes risk is not driven by a single biomarker but emerges from complex, stage-specific interactions between glycemic control, body composition, age, and lipid metabolism. While stratification improved insight into subgroup-specific risk patterns, its effectiveness was contingent on adequate sample size and class representation, underscoring the importance of balanced datasets in future work. Overall, these findings support the integration of stratified, interpretable machine learning models into clinical decision support systems to facilitate early detection, personalized intervention, and improved management of diabetes progression.

## DATA AVAILABILITY

Data will be available on requests.

